# Risk of COVID-19 related deaths for SARS-CoV-2 Omicron (B.1.1.529) compared with Delta (B.1.617.2)

**DOI:** 10.1101/2022.02.24.22271466

**Authors:** Isobel L. Ward, Charlotte Bermingham, Daniel Ayoubkhani, Owen J. Gethings, Koen B. Pouwels, Tomas Yates, Kamlesh Khunti, Julia Hippisley-Cox, Amitava Banerjee, Ann Sarah Walker, Vahé Nafilyan

## Abstract

**Objective:** To assess the risk of death involving COVID-19 following infection from Omicron (B.1.1.539/BA.1) relative to Delta (B.1.617.2).

**Design:** Retrospective cohort study.

**Setting:** England, UK, 1 December 2021 to 25 January 2022.

**Participants:** 1,035,163 people aged 18-100 years who tested positive for SARS-CoV-2 in the national surveillance programme, and had an infection identified as either Omicron- or Delta compatible.

**Main outcome measures:** Death involving COVID-19 as identified from death certification records. The exposure of interest was the SARS-CoV-2 variant identified from NHS Test and Trace PCR positive tests taken in the community (pillar 2) and analysed by Lighthouse laboratories. Cause-specific Cox proportional hazard regression models were adjusted for sex, age, vaccination status, previous infection, calendar time, ethnicity, Index of Multiple Deprivation rank, household deprivation, university degree, keyworker status, country of birth, main language, region, disability, and comorbidities. Additionally, we tested for interactions between variant and sex, age, vaccination status and comorbidities.

**Results:** The risk of death involving COVID-19 was 67% lower for Omicron compared to Delta and the reduction in the risk of death involving COVID-19 for Omicron compared to Delta was more pronounced in males than in females and in people under 70 years old than in people aged 70 years or over. Regardless of age, reduction of the risk of death from Omicron relative to Delta more was more pronounced in people who had received a booster than in those having received only two doses.

**Conclusions:** Our results support early work showing the relative reduction in severity of Omicron compared to Delta in terms of hospitalisation and extends this research to assess COVID-19 mortality. Our work also highlights the importance of the vaccination booster campaign, where the reduction in risk of death involving COVID-19 is most pronounced in individuals who had received a booster.

**What is already known on this topic:** The Omicron variant, which refers to the whole lineage (BA.1, BA.2, BA.3) had already been shown to be more transmissible than the Delta variant, but there is emerging evidence suggests that the risk of hospitalisation and risk of death within 28 days after a SARS-COV-2 test is lower. However, with a highly transmissible infection and high levels of population testing, definition of death within 28 days is more likely to be susceptible to misclassification bias due to asymptomatic or co-incidental infection. There is no study so far comparing the risk of COVID-19 death as identified from death certification records, with the cause of death assessed by the physician who attended the patient in the last illness.

**What this study adds:** Using data from a large cohort of COVID-19 infections that occurred in December 2021, we examined the difference in the risk COVID-19 death, as identified from death certification records, between the Delta and Omicron BA.1 variant. Our study shows that risk of death involving COVID-19 was reduced by 67% following infection with the Omicron BA.1 variant relative to the Delta variant after adjusting for a wide range of potential confounders, including vaccination status and comorbidities. Importantly, we found that the relative risk of COVID-19 mortality following Omicron versus Delta infection varied by age and sex, with lower relative risk in younger individuals and for males than females. The reduction in risk of death involving COVID-19 was also most pronounced in individuals who had received a booster.

## Introduction

On 27 November 2021 the UK Health Security Agency (UKHSA) identified the first UK cases of coronavirus disease 19 (COVID-19) variant B.1.1.529/BA.1, a variant of concern named, together with its sub-variants BA.2 and BA.3, as Omicron (Department of Health and Social Care, 2021). As the Omicron variant, which refers to the whole lineage (BA.1, BA.2, BA.3) had already been shown to be more transmissible, identifying whether the severity of disease, risk of hospitalisation, death or long-term complications is increased relative to Delta, is critical to enable pandemic and policy planning.

Omicron lineage B.1.1.529/BA.1 has a large number of mutations, 37 of which are in the Spike (S) protein (Ford, 2021), which leads to S-gene target failure (SGTF) in the molecular diagnostic assay (WHO, 2021). This can be identified from non-detectable S gene and a Cycle threshold (Ct) value of 30 or lower for the N and ORF1ab targets in positive polymerase chain reaction (PCR) tests using National testing data for England (based on the NHS Test and Trace programme), supplemented with data from the National Pathology Exchange (NPEx). Several studies have used a similar approach to compare the severity of Alpha (B.1.1.7) and Delta (B.1.617.2) with other variants [1]–[3].

Emerging data also indicate that risk of hospitalisation is lower following Omicron than Delta infection [4], [5], as is the risk of death within 28 days after a SARS-COV-2 test [5]. However, this analysis used death within 28 days of a positive test as a measure of COVID-19 death, rather than deaths involving COVID-19 identified using information from the death certificate, which include deaths at any time period and a cause of death classified by the physician who attended the patient in the last illness. Also, with a highly transmissible infection and high levels of population testing, definition of death within 28 days is more likely to be susceptible to misclassification bias due to asymptomatic co-incidental infection, than when infection rates are lower.

In this study, we compared the risk of COVID-19 death using death registration data in a large population-based cohort of people infected in England in December 2021, a period where both Delta and Omicron BA.1 variants were circulating, but Omicron BA.2 remained rare. In addition, we adjusted for a range of potential confounders, including pre-existing health conditions which previous work has not assessed.

## Methods

### Study data

We used data from the ONS Public Health Data Asset (PHDA), a linked dataset combining the 2011 Census, mortality records, the General Practice Extraction Service (GPES) data for pandemic planning and research, Hospital Episode Statistics (HES), NHS Test and Trace data (Pillar 2: swab testing for the virus in the wider population) and national vaccination data from the National Immunisation Management Service (NIMS). NIMS includes all vaccinations administered for all persons residing in England since the vaccination program started on 8^th^ Dec 2020.

To obtain NHS numbers, the 2011 Census was linked to the 2011-2013 NHS Participant Registers. Of the 53,483,502 Census records, 50,019,451 were linked deterministically. 555,291 additional matches were obtained using probabilistic matching (overall linkage rate: 94.6%). All subsequent linkages were conducted using NHS number. The ONS Public Health Data Asset include data on 35 million adults, an estimated 79% of the population of England in 2020.

### Study Population

The study population included all individuals who had a positive PCR test for COVID-19 between 1^st^ December 2021 and 31^st^ December 2021, reported as part of pillar 2 of NHS Test and Trace and analysed by Lighthouse Laboratories, who were enumerated at the 2011 Census and were living in England and were registered with a general practitioner on 1 November 2019. We specifically selected people who tested positive in December 2021 for our study population because both Delta and Omicron BA.1 variants were circulating during this period, but Omicron BA.2 remained rare. In January 2022, nearly all cases were due to the Omicron BA.1 or BA.2 variants, limiting the possibility to compare outcomes with Delta over the same period. Our sample contained 1,035,163 people who tested positive in the NHS Test and Trace pillar 2 with an Omicron BA.1- or Delta-compatible infection between 1^st^ and 31^st^ December 2021 and could be linked to the PHDA [Supplementary Table S1]. This covers 36.7% of all positive tests in England in December 2021.

Individuals entered the cohort on the index date which is the date of the first positive PCR test recorded between 1^st^ to 31^st^ December 2021. Individuals left the cohort on the earliest of: end of study date (25^th^ January 2022) (censored), death involving COVID-19 (event), or death from other cause (censored).

### Outcome

The primary outcome was time from positive PCR test to COVID-19 related death, defined as confirmed COVID-19 death identified by ICD-10 code U07.1 mentioned anywhere on the death certificate.

### Exposure

The exposure of interest was the COVID-19 variant in PCR positive tests taken in the community (pillar 2) and analysed by Lighthouse laboratories. Namely, defined by S-gene target failure (SGTF) as Omicron-compatible if S-negative, N-positive, ORF1ab-positive (with mean Ct <30 for N and ORF1ab) or Delta-compatible if S-positive/N-positive/ORF1ab-positive or ORF1ab-positive/S-positive or N-positive/S-positive, and average Ct <30. Of all Omicron- and Delta-compatible infections, a total small proportion (2.9%) of cases had mean Ct values greater than 30, indicative of a low viral load and were excluded because delta cases with high Ct values could be S-negative [see Supplementary Table S3]

### Covariates

Our main objective was to compare the risk of death involving COVID-19 between Delta and Omicron BA.1. We adjusted for a wide range of potential confounders of the relationship between variant type and the risk of COVID-19 death once infected, either in relation to vulnerability or testing behaviours, to account for any bias in our sample of individuals presenting as positive in the national surveillance programme.

Socio-demographic characteristics included age at time of infection (as a natural spline), sex, ethnicity (White/Black/South Asian/Other), region (North East, North West, Yorkshire and the Humber, East Midlands, West Midlands, East of England, London, South East, South West), disability, key worker status, Index of Multiple Deprivation rank (as a natural spline), country of birth (UK/Non-UK), university degree, household deprivation and English language ability. We also adjusted for baseline vaccination status (unvaccinated, one dose, two doses AstraZeneca ≤180 days previously, two doses mRNA vaccine (Pfizer or Moderna) ≤180 days previously, two doses AstraZeneca >180 days previously, two doses mRNA >180 days previously, any booster or third dose, which re refer to as boosters), previous infection (defined by a positive test at least 90 days before the date of the current positive test), for calendar date of infection using a natural spline, and for clinical risk factors by counting the number of conditions identified as being associated with an elevated risk of COVID-19 deaths in the QCovid 3 risk model (0 – 8). Further details of the chronic conditions are in Supplementary Table S2.

### Statistical Analysis

Characteristics of the study population were summarised overall, and stratified by variant type, using means for continuous variables and proportions for categorical variables.

We used cause-specific Cox proportional hazard regression model to estimate the hazard ratio of COVID-19 related death for individuals infected with Omicron versus Delta variants. For non-COVID-19 deaths, individuals were censored at the date of death if this occurred before the end of study date. We estimated four models, sequentially adjusted for age, sex, vaccination status and previous infection (Model 1); plus, calendar time (Model 2); plus, socio-economic factors (Model 3); and finally, plus pre-existing health conditions (Model 4).

To test whether the relative risk of mortality of Omicron varied by specific characteristics, we included interactions between variant type and age, and variant type and sex. In addition, we fitted separate fully adjusted models for aged 18-59, 60-69, 70+ years-olds to look at interactions between variant and vaccination status (unvaccinated, one dose, two doses and booster) an interaction between variant and health comorbidities (0, 1-2, 3+), grouped to have enough power to estimate the risk.

We assessed the proportional hazard assumption by testing for the independence between the scaled Schoenfeld residuals and time-at-risk. We used Schoenfeld residuals from the fitted Cox models, smoothed using generalized additive models, to assess whether relative differences in the hazard of COVID-19 death between variant was constant over time following the positive test.

## Results

### Characteristics of study population

There were 1,035,163 people in our study population. Of these, 814,011 (78.6%) individuals had Omicron-compatible and 221,152 (21.4%) Delta-compatible infections, with the number of Omicron infections increasing per day across the study period [Supplementary Figure S1]. This covers 36.7% of all positive tests in England in December 2021 [6]. In our study population, 54% of infections were in females [Table 1, Supplementary Table S3]. The mean age at infection was two years younger in those infected with Omicron (39.9 years, SD=15.2) than Delta (42.2 years, SD=13.1). There were 128 deaths involving to COVID-19 and 53 deaths not involving COVID-19 in those infected with Omicron, and 189 and 28, respectively, in those infected with Delta [Table 2]. The mean time from positive result to COVID-19 related death was 13 days (SD=7.0) for Omicron and 16 days (SD=9.6) for Delta.

**Table 1.** Baseline characteristics of patients infected with either Omicron or Delta variants

| Variable | Group | Total | Delta (% of total in group) | Omicron (% of total in group) |
| --- | --- | --- | --- | --- |
| Country of birth | Non-UK | 117764 | 21.3 | 78.7 |
|  | UK | 917399 | 21.4 | 78.6 |
| University degree | No | 788972 | 20.0 | 80.0 |
|  | Yes | 246191 | 25.6 | 74.4 |
| Disability | None/Day-to-day activities not limited or limited a little | 1015955 | 21.3 | 78.7 |
|  | Day-to-day activities limited a lot | 19208 | 23.2 | 76.8 |
| Ethnicity | Black | 39305 | 11.9 | 88.1 |
|  | Other | 63944 | 16.3 | 83.7 |
|  | South Asian | 46034 | 19.9 | 80.1 |
|  | White | 885880 | 22.2 | 77.8 |
| Household deprivation | Household is not deprived in any dimension | 607764 | 21.5 | 78.5 |
|  | Household is deprived in 1 dimension | 280532 | 21.0 | 79.0 |
|  | Household is deprived in 2 dimensions | 103553 | 21.8 | 78.2 |
|  | Household is deprived in 3 dimensions | 28721 | 23.2 | 76.8 |
|  | Household is deprived in 4 dimensions | 2558 | 24.9 | 75.1 |
|  | Missing | 12035 | 15.2 | 84.8 |
| Key worker † | No | 253011 | 23.8 | 76.2 |
|  | Yes | 782152 | 20.6 | 79.4 |
| Main Language | English | 66908 | 22.7 | 77.3 |
|  | Non-English | 968255 | 21.3 | 78.7 |
| Previous COVID-19 infection | No | 979310 | 22.3 | 77.7 |
|  | Yes | 55853 | 4.1 | 95.9 |
| Region | North East | 46624 | 18.9 | 81.1 |
|  | North West | 192220 | 19.2 | 80.8 |
|  | Yorkshire and the Humber | 120802 | 23.3 | 76.7 |
|  | East Midlands | 83255 | 24.4 | 75.6 |
|  | West Midlands | 91292 | 27.8 | 72.2 |
|  | East of England | 118094 | 24.9 | 75.1 |
|  | London | 188942 | 14.1 | 85.9 |
|  | South East | 155301 | 21.8 | 78.2 |
|  | South West | 38633 | 30.5 | 69.5 |
| Sex | Male | 478276 | 21.2 | 78.8 |
|  | Female | 556887 | 21.5 | 78.5 |
| Count of comorbidities ‡ | 0 | 908654 | 21.5 | 78.5 |
|  | 1-2 | 122586 | 20.1 | 79.9 |
|  | 3 | 3923 | 20.2 | 79.8 |
| Vaccination status | Booster | 233035 | 8.8 | 91.2 |
| Vaccination status | One dose | 32790 | 26.0 | 74.0 |
| Vaccination status | Two doses AZ > 180 days | 86238 | 24.2 | 75.8 |
| Vaccination status | Two doses AZ ≤ 180 days | 38385 | 37.8 | 62.2 |
| Vaccination status | Two doses mRNA > 180 days | 171850 | 30.1 | 69.9 |
| Vaccination status | Two doses mRNA ≤ 180 days | 359755 | 19.7 | 80.3 |

† Information on Census 2011 variables that were used to define key worker status. ‡ Count of comorbidities grouped for disclosure control reasons, added as linear continuous predictor to fully adjusted model (Model 4).

**Table 2.** Counts of cases, deaths involving COVID-19 and not involving COVID-19

|  | **Total** | **Omicron** | **Delta** |
| --- | --- | --- | --- |
| Positive COVID-19 cases | 1035163 | 814011 | 221152 |
| Deaths involving COVID-19 | 317 | 128 | 189 |
| *Age 18-59 years* | 50 | 42 | 8 |
| *Age 60-69 years* | 52 | 42 | 10 |
| *Age 70+ years* | 215 | 105 | 110 |
| Deaths not-involving COVID-19 | 81 | 53 | 28 |

### Difference in risk of COVID-19 mortality by variant

The risk of death involving COVID-19 was 67% lower (HR=0.33, 95%CI: 0.24, 0.45) [Table S4] for Omicron compared to Delta infections in our fully adjusted model (Model 4), accounting for sex, age, vaccination status, previous infection, calendar time, ethnicity, Index of Multiple Deprivation rank, household deprivation, university degree, keyworker status, country of birth, main language, region, disability, and health risk factors defined in the QCovid 3 model [Figure 1]. In our minimally adjusted model (Model 1) accounting for sex, vaccination status, age and previous infection, the risk of death was 78% lower (HR=0.22, 95%CI: 0.17, 0.28) for Omicron versus Delta. Whilst also adjusting for the date of infection (Model 2) reduced the difference somewhat (HR=0.31, 95%CI: 0.23, 0.43), further adjusting for socio-demographic characteristics (Model 3) and pre-existing health conditions (Model 4) had little impact on the relative difference between Omicron and Delta related mortality (HR=0.32 and 0.33 respectively).

**Figure 1.**
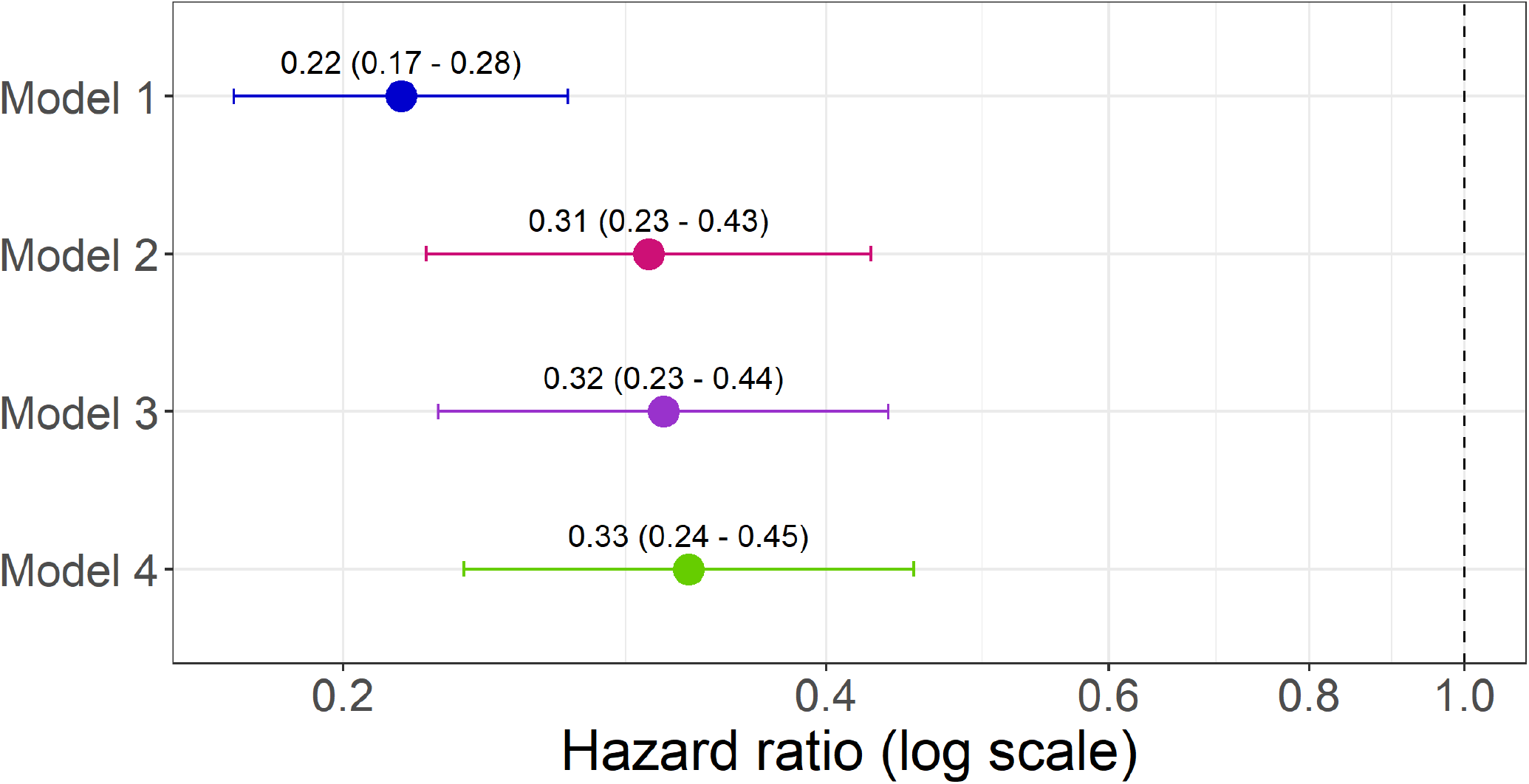
Hazard ratio for death involving COVID-19 death for Omicron vs Delta infections for fully adjusted (Model 4) and alternative models. The risk is shown for Omicron relative to Delta, with the dashed line showing HR = 1. Footnote: **Model 1** adjusted for sex, age (natural spline), vaccination status and previous infection. **Model 2** adjusted for sex, age (natural spline), vaccination status, previous infection, and calendar time (natural spline). **Model 3** adjusted for sex, age (natural spline), vaccination status, previous infection, calendar time, ethnicity, Index of Multiple Deprivation rank (natural spline), household deprivation, university degree, keyworker status, country of birth, main language, region and disability. **Model 4** adjusted for sex, age (natural spline), vaccination status, previous infection, calendar time, ethnicity, Index of Multiple Deprivation rank (natural spline), household deprivation, university degree, keyworker status, country of birth, main language, region, disability, and comorbidities.

### Difference in risk of death involving COVID-19 by variant and age and sex

Estimates of the difference in the relative risk of death involving COVID-19 between Omicron and Delta by sex and age are presented in Figure 2. The difference in mortality risk varied strongly by age (X^2^(4) = 22.8, p < 0.001) with greater reduction in COVID-19 mortality with Omicron compared to Delta for people under aged 18-59 (HR=0.13, 95%CI: 0.06, 0.30) and 60-69 years (HR=0.14, 95%CI: 0.05, 0.36) than aged 70 or over (HR=0.45, 95%CI: 0.16, 1.24, heterogeneity p-value = < 0.001). For the interaction between sex and variant (X^2^(1) = 5.65, p < 0.017), the reduction in mortality risk for COVID-19 is more pronounced in males (HR=0.25, 95%CI: 0.17, 0.37) than in females (HR=0.44, 95%CI: 0.28, 0.68, heterogeneity p-value = 0.016).

**Figure 2.**
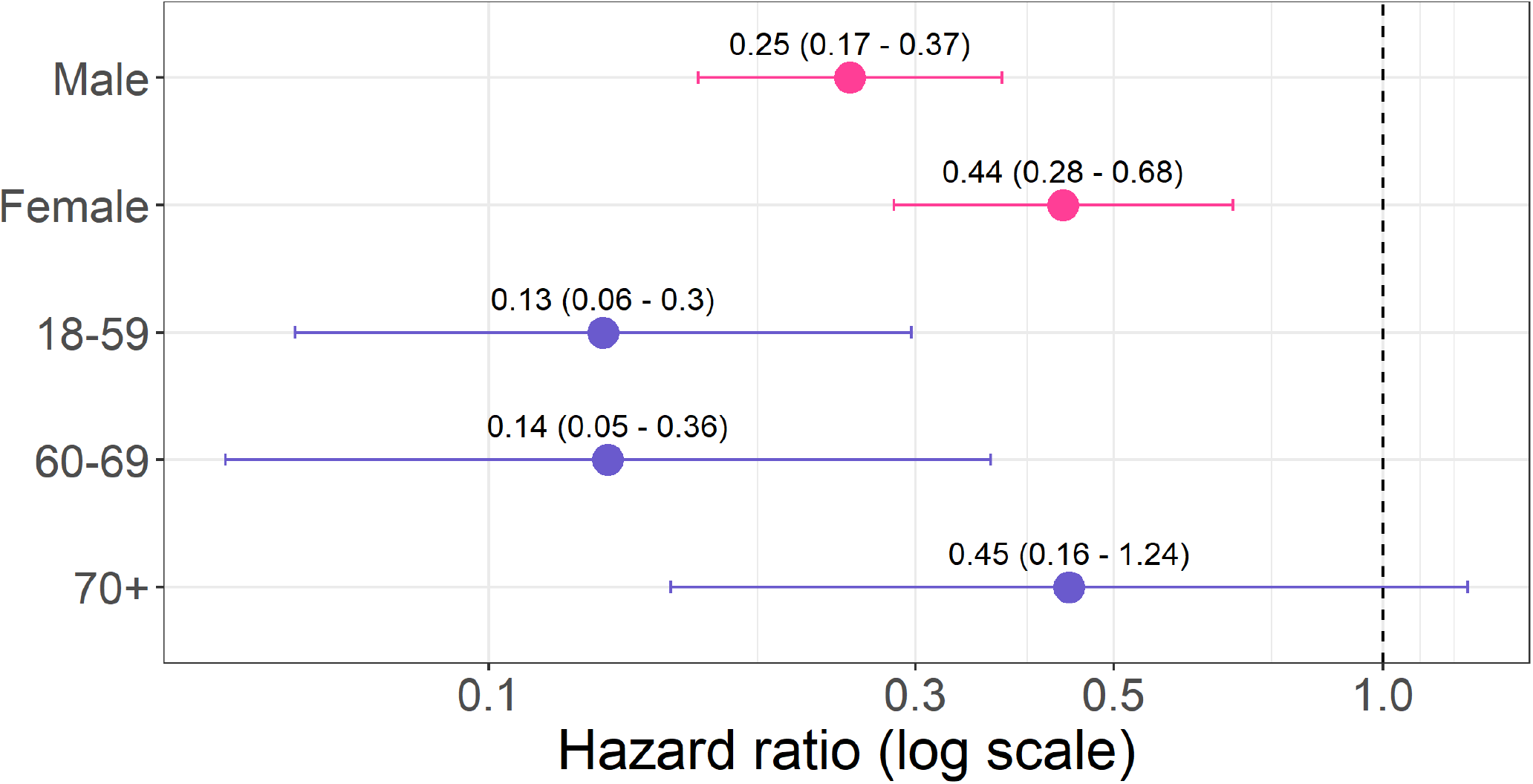
Hazard ratio for death involving COVID-19 for Omicron vs Delta infections by sex and age. The risk is shown for Omicron relative to Delta, with the dashed line showing HR = 1. To look at the interaction between variant type and sex, the model was fully adjusted was run (Model 4) with an interaction term for variant and sex. For the variant type and age, the fully adjusted model also included a variable flagging age group 18-59, 60-69 or 70+ which was interacted with variant.

### Difference in risk of death involving COVID-19 by variant and vaccination status and comorbidities

To assess whether the difference in risk of death between Omicron and Delta varied by age and comorbidities, we fitted fully adjusted models separately for different ages (18-59, 60-69, 70+ years-old) and tested for an interaction between variant and vaccination status, and separately between variant and number of health comorbidities within each age group [Figure 3]. To assess the interaction between variant and vaccination, vaccination status was re-grouped to unvaccinated, one dose, two doses and booster. It was not possible to compute accurate confidence intervals for risk in the ‘one dose’ group due to lack of power. Regardless of age, having received a booster (18-59: HR=0.06, 95%CI: 0.01, 0.25; 60-69: HR=0.06, 95%CI: 0.01; 70+: HR=0.33, 95%CI: 0.10, 1.06) reduced the risk of mortality from Omicron relative to Delta more compared to having received only two doses (18-59: HR=0.56, 95%CI: 0.13, 2.45; 60-69: HR=0.22, 95%CI: 0.06, 0.88; 70+: HR=0.99, 95%CI: 0.31, 3.10).

**Figure 3.**
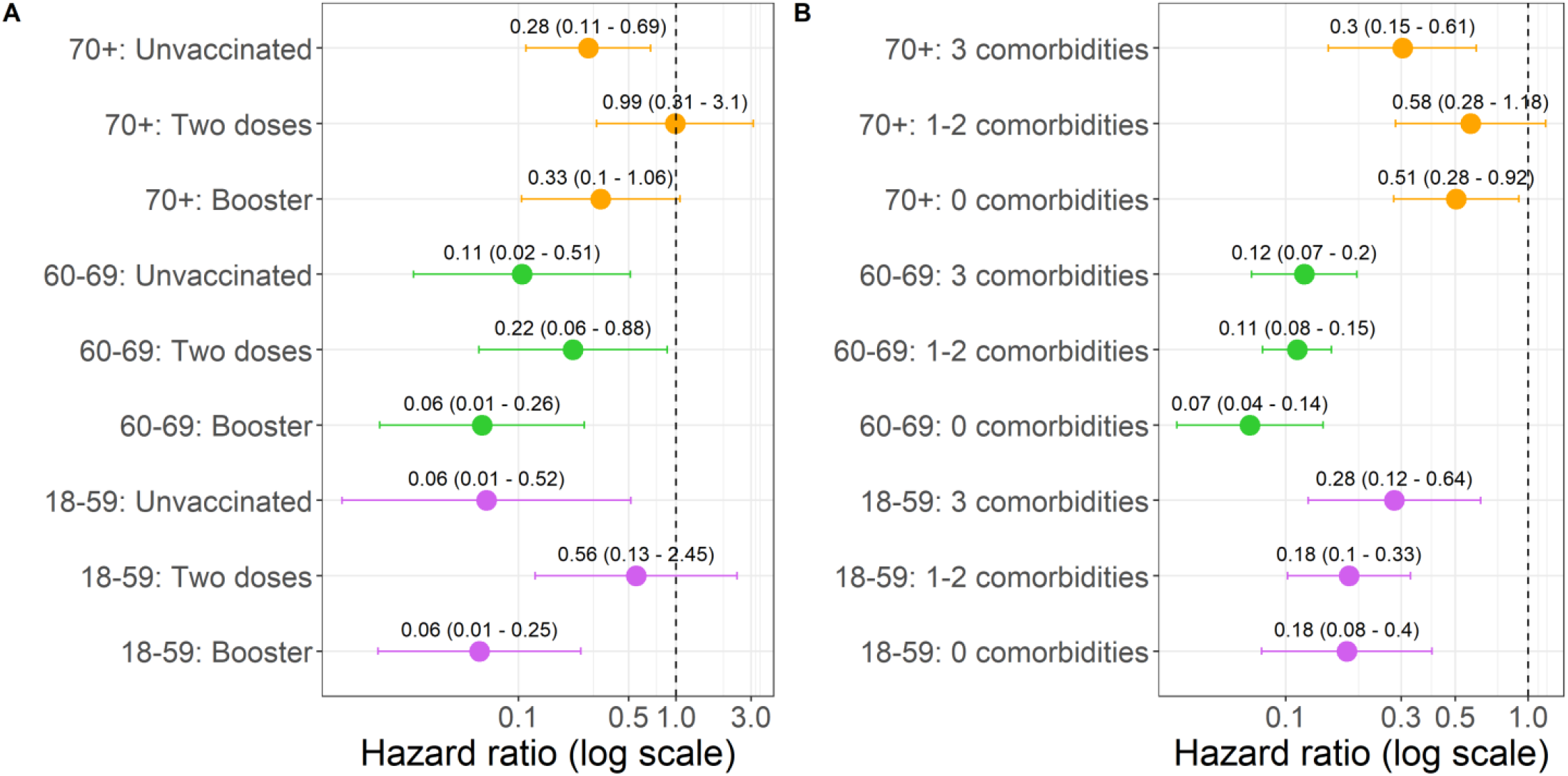
Hazard ratio for death involving COVID-19 death for Omicron vs Delta infections for variant vaccination status and variant comorbidities **A**. The fully adjusted model was used to assess interactions between variant and vaccination status and comorbidities separately for age 18-59, 60-69 and 70+ year olds. Vaccination status was re-grouped to unvaccinated, one dose (not plotted), two doses and booster. **B**. Comorbidities were grouped to flag a total of 0, 1-2 of 3+.

The risk of COVID-19 related death following Omicron infection was lower than Delta, for all age groups, regardless of the number of comorbidities (grouped to 0, 1-2 and 3+), but there was no evidence that the relative mortality risk varied by the number of comorbidities [Figure 3].

We tested the proportional hazard assumption by testing for the independence between the scaled Schoenfeld residuals and time-at-risk. The test failed to reject the independence for the exposure (p = 0.07), suggesting that the proportional hazard assumption was unlikely to be violated.

## Discussion

### Main Findings

Using data from a large cohort of COVID-19 infections that occurred in December 2021, we examined the difference in mortality between the Delta and Omicron BA.1 variant. Our study shows that risk of death involving COVID-19 was reduced by 67% following infection with the Omicron BA.1 variant relative to the Delta variant after adjusting for a wide range of potential confounders, including vaccination status and comorbidities. Importantly, we found that the relative risk of COVID-19 mortality following Omicron versus Delta infection varies by age, with lower relative risk in younger individuals.

### Comparison with other studies

Early work exploring the clinical severity of COVID-19 Omicron variant in a South African cohort found significantly reduced odds of hospitalisation following SGTF versus non-SGTF infection across the same period [4]. A subsequent study in California on positive PCR tests between 30 November 2021 and 1 January 2022 also showed risk reductions for hospital admission, ICU admission and mortality following Omicron relative to Delta infections [7]. In Canada, in a matched sample, the risk of hospitalisation or death was found to be 65% lower among Omicron than Delta cases [8].

Our results extend these initial analyses quantifying intrinsic risk of Omicron severity in terms of hospital admissions, to COVID-19 mortality. Nyberg et al. 2022 report a reduction in death following Omicron infection (HR=0.31) relative to Delta, which is similar to our findings. Importantly, our results account for more sociodemographic factors and comorbidities, and highlight that the reduction in risk remains consistent even after adjusting for these additional variables. Furthermore, our study specifically quantifies the risk of cause-specific COVID-19 mortality, utilising death registration data, unlike previous work which has defined COVID-19 death as death within 28 days of a positive COVID-19 test, resulting in large

Given the emergence of the Omicron variant resulted in an increased rate of transmission, the number of Omicron cases in our sample of infected individuals increased significantly across the study period. To account for the difference in infection rate across the period, a cubic spline for calendar time was included in Models 2 – 4. The BA.2 sub-variant of Omicron does not have the spike gene deletion that causes SGTF. The UK noted an increase in the number of sub-variant BA.2 cases in the week commencing 3^rd^ January 2022 [9]. Our data include Omicron-compatible and Delta-compatible infections identified between 1^st^ and 31^st^ December 2021, therefore in a period where BA.1 was prominent, and Omicron could be identified from SGTF.

These results provide clear evidence that the risk of COVID-19 mortality following infection with Omicron is significantly less than Delta in the UK.

## Strengths and limitations

First, we use a large sample of positive cases from the national testing programme, allowing to precisely estimate the relative risk of COVID-19 death following infection with Omicron BA.1 and Delta. Second, by linking these infection data to information on vaccination status, comprehensive socio-demographic characteristics from the Census and information on pre-existing conditions based on primary care and hospital data, we were able to estimate the difference in mortality between the Omicron BA.1 and Delta variants, adjusting for a wide range of potential confounders, including vaccination status with manufacturer type, and key worker status. We also tested for the relative mortality risk of Omicron depended on the vaccination status, by including interactions between variant type and vaccination status. Third, we use death certificate data to confirm COVID-19 mortality, preventing our sample being conflated with non-COVID-19 related deaths of individuals that die of other causes following a positive COVID-19 test. We also compared the outcomes during the same time periods overcoming any differences due to changes in management of infected patients over the time period of the pandemic.

The main study limitation is an ascertainment bias since the data do not cover all SARS-CoV-2 infections, but only a subset of people who tested positive as part of the national testing programme in the community and analysed by Lighthouse laboratories. Tests conducted in the community but processed by other laboratories and tests conducted in hospitals could not be used because they do not use the S-gene molecular diagnostic assay, which we used to identify the variant type. A limitation of our work is not having access data to derive COVID-19 variants from tests in hospital (NHS Pillar 1), and explains why our total sample is smaller than other research [5]. Differences in testing behaviours between groups may bias the estimates of risk of COVID-19 death among people who tested positive. If some people only get tested if they experience severe symptoms, the estimated risk of death would be higher in this group than in people who get tested more routinely, even if the population has the same underlying risk. To mitigate this issue, we also adjusted the models for factors that may affect the propensity to get tested and may also be related to the severity of a SARS-CoV-2 infection, including ethnicity, region, calendar date of infection, and key worker status. Socio-demographic information was used from Census 2011 as was the most up to date at time of publishing, however future validation work should be conducted once Census 2021 data has been released and potentially using more granular breakdowns of variables, such as region.

Because of death registration delays, not all deaths that occurred in the period may yet have been registered. Deaths that occurred amongst people who tested positive in late December are less likely to have been registered than those which occurred in people who tested positive at the beginning of the month. As the proportion of cases which are from the Omicron variant increased during December, the delay in death registration, if unaccounted for, could lead to underestimation of the severity of the Omicron variant. However, we accounted for the effect of registration delay in December by adjusting for calendar time of infection in our models, reducing the difference between Omicron compared to Delta as expected.

## Conclusions

Given the emergence of the more transmissible Omicron BA.1 variant, there was an urgent healthcare requirement to quantify the risk of COVID-19 death relative to other variants to support pandemic planning responses. Our results support early work showing the relative reduction in severity of Omicron compared to Delta in terms of hospitalisation and extends this research to assess COVID-19 mortality, being the first to our knowledge to assess cause-specific COVID-19 death using death certification to accurately capture COVID-19 deaths. Our work also highlights the importance of the vaccination booster campaign, where the reduction in risk of death involving COVID-19 is most pronounced in individuals who had received a booster. However, mortality is only one metric that should be considered when assessing of the impact of COVID-19 and subsequent work should investigate long-term outcomes of infection, such as the prevalence of long COVID following Omicron infection relative to Delta.

## Data Availability

Information on data availability and access is available via the Secure Research Service: https://www.ons.gov.uk/aboutus/whatwedo/statistics/requestingstatistics/approvedresearcherscheme

## Acknowledgements

TY received funding from a grant from the UKRI (MRC)-DHSC (NIHR) COVID-19 Rapid Response Rolling Call (MR/V020536/1) and is part of the Data and Connectivity National Core Study, led by Health Data Research UK in partnership with the Office for National Statistics and funded by UK Research and Innovation (HDRUK2020.138)

ASW is an NIHR Senior Investigator and is supported by the Oxford Biomedical Research Centre. The views expressed are those of the authors and not necessarily those of the NHS, the NIHR, UKSHA or the Department of Health and Social Care. KBP is supported by the Huo Family Foundation.

## Ethical approval

Ethical approval was obtained from the National Statistician’s Data Ethics Advisory Committee (NSDEC(20)12).

## Funding

None

## Contributors

IW, ASW and VN conceptualised and designed the study. IW and CB prepared the study data. IW performed the statistical analysis, which were quality checked by CB and VN. All authors contributed to interpretation of the findings. IW and VN wrote the original draft. All authors contributed to review and editing of the manuscript and approved the final manuscript.

## Supplementary Tables

**Table S1:** Sample flow

| Processing Stage | Count |
| --- | --- |
| Total number of records in Test and Trace | 382458741 |
| Sample that links to Census/PHDA | 299084765 |
| Sample with non-logical dates removed | 297701958 |
| Sample with positive infection | 10647556 |
| Sample with PCR test | 9140824 |
| Sample with Pillar 2 PCR | 7634416 |
| Sample tested in Lighthouse Laboratory | 5245792 |
| De-duplication of records | 5238805 |
| Sample with one infection per person per 90-day spell | 5092528 |
| Sample with infection date ≥ 01-12-2021 | 1339606 |
| Removal of erroneous vaccination dates | 1339326 |
| Individuals aged 18-100 on infection date | 1200181 |
| Removal of erroneous date of death | 1200165 |
| Sample with Omicron- or Delta-compatible infection | 1067018 |
| Exclusion of non-England region from Census records | 1066587 |
| Exclude Omicron/Delta compatible infections with Ct average $>$ 30 | 1035163 |

**Table S2:** Variables grouping

| Variable | Group |
| --- | --- |
| Key worker † | Health professionals |
|  | Health associate professionals |
|  | Support staff |
|  | Social care |
|  | Education |
|  | Food retail and distribution |
|  | Taxi and cab drivers and chauffeur |
|  | Bus and coach drivers |
|  | Van drivers |
|  | Other transport workers |
|  | Police and protective services |
|  | Sanitary Workers |
| Comorbidity | Asthma |
|  | Atrial fibrillation |
|  | Blood or bone marrow cancer |
|  | Chronic kidney disease |
|  | Congenital heart problems |
|  | COPD |
|  | Coronary heart disease |
|  | Cystic fibrosis |
|  | Dementia |
|  | Diabetes |
|  | Epilepsy |
|  | Heart failure |
|  | Learning disability or Downs Syndrome |
|  | Liver cirrhosis |
|  | Lung or oral cancer |
|  | Motor neurone disease |
|  | Multiple sclerosis |
|  | Myasthenia |
|  | Huntington's disease |
|  | Chorea |
|  | Parkinson's disease |
|  | Peripheral vascular disease |
|  | Prior fracture of hip, wrist, spine, humorous |
|  | Pulmonary hypertension or fibrosis |
|  | Rheumatoid arthritis |
|  | Systemic lupus erythematosus |
|  | Severe combined immunodeficiency |
|  | Sickle cell |
|  | Stroke |
|  | Transient ischaemic attack |
|  | Thrombosis |
|  | Pulmonary embolism |
|  | Schizophrenia |

† Key workers status is defined based on the occupation and industry information collected at 2011 Census and includes people working in education & childcare, food & necessity goods, health & social care, public services, national & local government, public safety & national security, transport, utilities & communication

**Table S3:** Continuous variables in model average/SD

| Variable | Average (SD) |
| --- | --- |
| Age | 40.39 (14.82) |
| IMD rank | 17098.29 (9394.9) |
| Calendar time | 19.92 (7.38) |

**Table S4:** Risk of mortality from COVID-19 cases with Omicron compared to Delta for each model. ***Model 1*** *adjusted for sex, age (natural spline), vaccination status and previous infection*. ***Model 2*** *adjusted for sex, age (natural spline), vaccination status, previous infection, and calendar time (natural spline)*. ***Model 3*** *adjusted for sex, age (natural spline), vaccination status, previous infection, calendar time, ethnicity, Index of Multiple Deprivation rank (natural spline), household deprivation, university degree, keyworker status, country of birth, main language, region and disability*. ***Model 4*** *adjusted for sex, age (natural spline), vaccination status, previous infection, calendar time, ethnicity, Index of Multiple Deprivation rank (natural spline), household deprivation, university degree, keyworker status, country of birth, main language, region, disability, and comorbidities*.

|  | HR | CI |
| --- | --- | --- |
| Model 1 | 0.22 | 0.17-0.28 |
| Model 2 | 0.31 | 0.23-0.43 |
| Model 3 | 0.32 | 0.23-0.44 |
| Model 4 | 0.33 | 0.24-0.45 |

**Figure S1.**
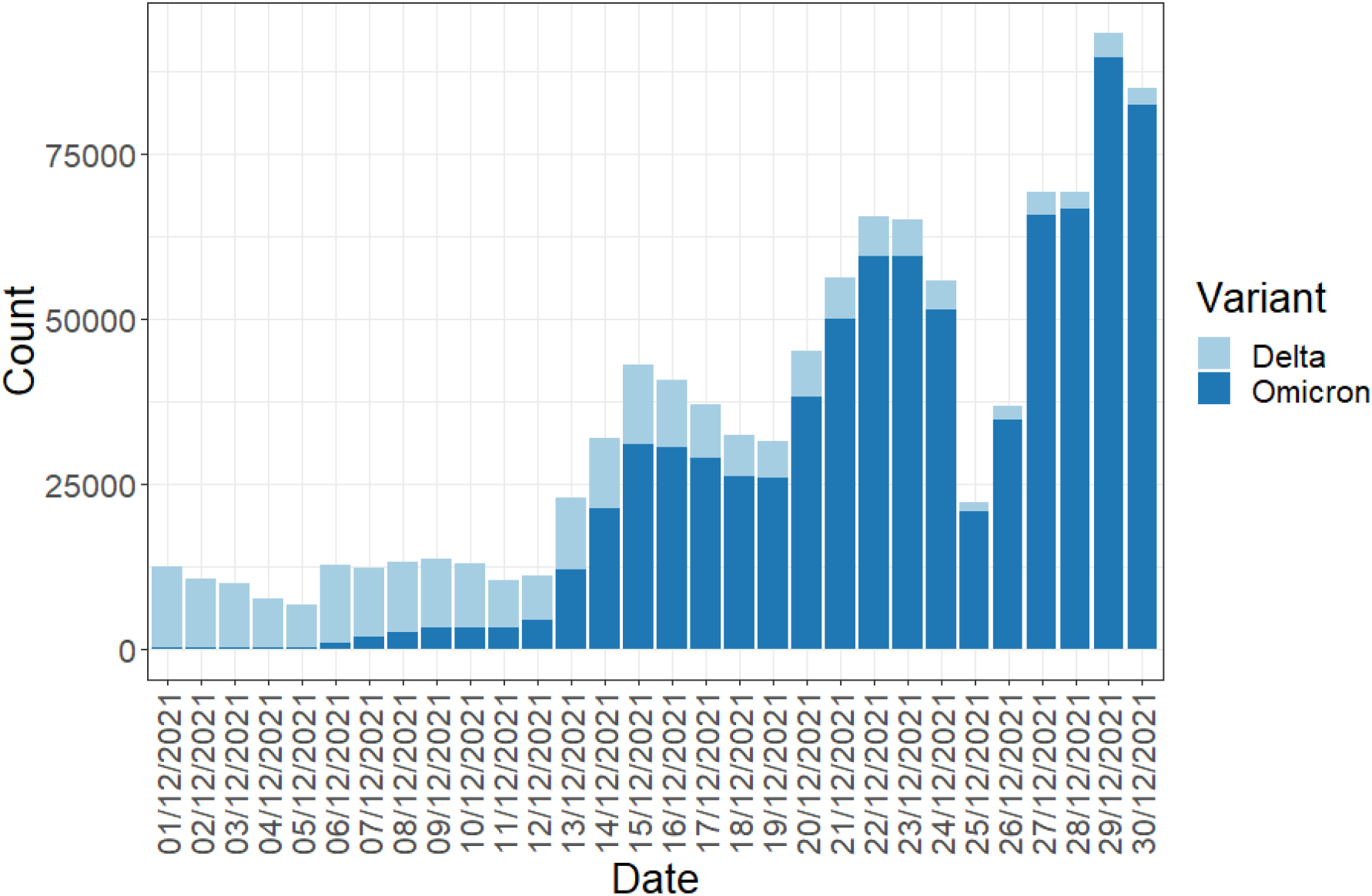
Number of infections by variant The stacked bar plot shows the number of infections by variant per day between 1^st^ December 2021 and 31^st^ December 2021. The date is the date of specimen from NHS Test and Trace. Omicron infections are shown in dark blue, and delta in light blue.

## Notes

### Competing Interest Statement

The authors have declared no competing interest.

### Funding Statement

No funding

## Bibliography

[1] K. A. Twohig et al., “Hospital admission and emergency care attendance risk for SARS-CoV-2 delta (B.1.617.2) compared with alpha (B.1.1.7) variants of concern: a cohort study,” Lancet Infect. Dis., vol. 22, no. 1, pp. 35–42, 2022, doi: 10.1016/S1473-3099(21)00475-8.

[2] M. Patone et al., “Mortality and critical care unit admission associated with the SARS-CoV-2 lineage B.1.1.7 in England: an observational cohort study,” Lancet Infect. Dis., vol. 21, no. 11, pp. 1518– 1528, Nov. 2021, doi: 10.1016/s1473-3099(21)00318-2.

[3] N. G. Davies et al., “Increased mortality in community-tested cases of SARS-CoV-2 lineage B.1.1.7,” Nature, vol. 593, no. 7858, pp. 270–274, 2021, doi: 10.1038/s41586-021-03426-1.

[4] N. Wolter et al., “Early assessment of the clinical severity of the SARS-CoV-2 omicron variant in South Africa: a data linkage study,” Lancet, vol. 399, no. 10323, pp. 437–446, 2022, doi: 10.1016/S0140-6736(22)00017-4.

[5] T. Nyberg et al., “Comparative analysis of the risks of hospitalisation and death associated with SARS-CoV-2 Omicron (B.1.1.529) and Delta (B.1.617.2) variants in England,” 2022.

[6] UK Government, “UK Coronavirus Dashboard,” 2022. https://coronavirus.data.gov.uk/details/cases (accessed Feb. 22, 2022).

[7] L. Joseph, V. Hong, M. Patel, R. Kahn, M. Lipsitch, and S. Tartof, “Clinical outcomes among patients infected with Omicron (B.1.1.529) SARS-CoV-2 variant in southern California,” medRxiv, no. 165, pp. 1–13, 2021.

[8] A. C. Ulloa, S. A. Buchan, N. Daneman, and K. A. Brown, “Early estimates of SARS-CoV-2 Omicron variant severity based on a matched cohort study, Ontario, Canada,” medRxiv, p. 2021.12.24.21268382, 2022, [Online]. Available: https://www.medrxiv.org/content/10.1101/2021.12.24.21268382v2 %0A https://www.medrxiv.org/content/10.1101/2021.12.24.21268382v2.abstract.

[9] UKHSA, “SARS-CoV-2 variants of concern and variants under investigation in England Technical briefing 34,” no. January, 2022.

